# Prevalence and determinants of food insecurity among households headed by mothers who are female sex workers in three African Countries: Validation and application of food insecurity experience scale

**DOI:** 10.1101/2024.06.21.24309302

**Authors:** Swarna D.S. Weerasinghe, Jennifer A. Jackson, Diane D. Stadler, Wendy L. Macias-Konstantopoulos, Brian Wills

## Abstract

Reduction in prevalence of food insecurity, a key factor in reaching the United Nations Sustainable Development Goal 2.2, was challenged by COVID-19 lockdowns in low- and middle-income countries, which impacted the affordability and accessibility of food. Prevalence and determinants of food insecurity among female sex workers who are mothers (FSWM) and heads of households (FSWM-HH) in the African region have not been previously assessed. This study validated the food insecurity experience scale (FIES) using data collected from 852 FSWM-HH living in the Democratic Republic of the Congo (DRC), Kenya, or Nigeria. Results were calibrated on the global metric to compare with the country-level prevalence of household severe food insecurity (HSFI). Associated individual and household determinants and their intersections were examined to identify vulnerable sub-groups. The FIES reliably assessed (fit index=0.75) household food insecurity among FSWM-HH using a 7-item scale, omitting the item of being worried about not having enough food to eat. The prevalence of HSFI was 59.6% in the DRC, 77.8% in Kenya, and 89.0% in Nigeria and compared to FAO 2016-2018 country levels, 1.5, 3.0, and 4.5 times higher, respectively, than previous estimates. Determinants of HSFI among FSWM-HH were similar to the African region, except FSWM-HH composition of children, but the magnitudes of odds were higher. Multiple logistic regression revealed important ecological relationships. FSWM-HH with 0-3 children living with them (AOR=1.63,95% CI:1.13-2.37, reference ≥4 children) and mothers having no schooling or primary education (AOR=1.92,95% CI:1.33-2.78, reference with higher education) significantly determined HSFI. Prevalence of HSFI was highest among two subgroups of FSWM-HH: those ≥ 30 years of age, without partners (93.7%, p=0.002) and those with low levels of education with 0-3 children living with them (85%, p=0.03). Food programs and welfare policies may be effective strategies to reduce HSFI among this high-risk group of mothers with children.

## Introduction

Food insecurity, a global crisis, was exacerbated by pandemic-related lockdowns and disruption to human economic activities, which disproportionately affected vulnerable groups of individuals and households in low- and middle-income countries (LMIC) [1]. Escalating rates of undernourishment in all regions of Africa rose to 22.7% in 2016, an increase of 2.7% from 2015, leaving more than 200 million people severely food insecure, even before the pandemic [2].

Over one-fifth of the global population in the African region experienced hunger in 2020, leaving 346.4 million suffering from severe food insecurity and an additional 452 million suffering from moderate food insecurity, further widening the gap to achieving the Sustainable Development Goal (SDG) 2.2 targets of ending malnutrition and hunger by 2030 [3]. National and international collaboration is identified as a key strategy to identify and address challenges to achieving food security [3]. To meet the SDG 2.2 goal and make concerted efforts toward combating this public health crisis, countries must focus attention on their most vulnerable populations. Research-based evidence is needed to identify the impact, understand the determinants, and estimate the prevalence and severity of food insecurity in these populations.

Between 2018 and 2020, the three countries in Africa with the highest level of moderate to severe food insecurity were Kenya (68%), Nigeria (58%), and the Democratic Republic of the Congo (DRC) (70%) [3]. Within each country, female sex workers who are mothers and who are heads of households (FSWM-HH) and members living in the same household, including children, are among those at the highest risk of experiencing severe food insecurity. Our study explored household food insecurity experiences, using data collected from FSWM-HH in these three Sub-Saharan African countries, and examined the validity of the Food and Agriculture Organization (FAO) Food Insecurity Experience Scale (FIES) measurement tool application to this data, to uncover determinants of household severe food insecurity (HSFI) and to identify the demographically determined subgroups of FSWM-HH. This study’s goal was to identify demographically characterized subgroups critical to meeting SDG 2.2 specific to the African region [2, 3] to ensure access to food to all by 2030, specifically the extremely vulnerable populations of FSWM-HH living in three African countries with the highest rates of malnutrition and food insecurity.

The sub-Saharan region of Africa is entangled with poverty, food insecurity, and HIV/AIDS [2, 5]. Research carried out in this region has shown that hunger and malnutrition push women to engage in the sex industry [5]. In Nigeria, 35% of a study sample indicated their reason for becoming a sex worker was poverty and lack of money to purchase food [6]. The same researchers observed a significant relationship between poverty, food insecurity, HIV/AIDS, and unwanted pregnancies in this population, a relationship mediated by low rates of pregnancy prevention measures (75.6%) [6]. The researchers also identified the reduction of poverty and food insecurity as a means to control HIV/AIDS [6]. A systematic review of 10 peer-reviewed articles on female sex workers (FSW) and HIV treatment and care in sub-Saharan Africa revealed that sub-optimal treatment outcomes are partly due to poor nutrition and food insecurity [7]. Women-led households are known to have higher rates of severe food insecurity than men-led households due to the poor economic status of these women [8]. Thus, exploring household food insecurity among FSWM-HH in the three Sub-Saharan countries of Kenya, Nigeria, and the DRC provides noteworthy insight into ways to alleviate hunger, malnutrition, and other related public health concerns.

One in 20 women in Sub-Saharan Africa have reportedly engaged in sex for money, and Kenya and Nigeria were noted to have the highest percentages of adult women engaging in sex work for money [9]. Literature suggests special consideration be given to vulnerable women since food insecurity can drive them to engage in high-risk sexual behaviors,[8] which in turn can exacerbate their vulnerability to food insecurity. Despite government efforts to provide anti-retroviral treatment (ART) to FSW, food insecurity has been linked to sub-optimal outcomes in engaging in HIV treatment and care [7].

There is ample evidence linking the COVID-19 pandemic to food insecurity in the Sub-Saharan region and the augmented effect on vulnerable women and FSW [10]. A study conducted among poor urban residents in Malawi, including sex workers, indicated they had little or no money to buy healthy food due to their loss of income resulting from the lockdown [11]. A cross-sectional survey conducted in Nigeria in September 2021 revealed 2.47 higher odds of food insecurity among FSW compared to those women who identified as transgender or living with a disability [10]. The estimated prevalence of food insecurity among the study sample was 76.1% [10].

Our theoretical underpinning of FSWM household severe food insecurity (HSFI) exploration is governed by the United Nations conceptual framework of social determinants of health (SDH) that was later adapted to understand the adverse health outcomes such as food insecurity [12] resulting from economic volatility induced by the pandemic among TB patients in South Africa [12]. Some disadvantaged groups, such as FSWM and their families, living in the same region are equally or more vulnerable to food insecurity. The study sample of FSWM-HH referenced in this article is at an added disadvantage due to multiple social vulnerabilities, [13] including living in an LMIC, headed by a woman, a mother, and a sex worker. Within the SDH framework, we used an intersectionality lens to examine the impact of the individual FSWM and household intersecting demographic characteristics to identify the most vulnerable subgroups for severe food insecurity.

We used the Food Insecurity Experience Scale (FIES) tool developed by the Voices of the Hungry project, launched by the FAO of the United Nations in 2012. The FIES was piloted in four sub-Saharan African countries, Angola, Ethiopia, Malawi, and Nigeria. Linguistic adaptations were made before release in 2014, and the revised tool was later validated in 151 countries [14, 15], including the three study countries. Although the FIES was not validated for use among FSWM-HH, it was applied to collect data from a nationally representative sample of individuals living in the three countries represented in this study. Though the psychometric properties of the FIES have been extensively validated among 151 country sample gathered from global study populations, the applicability of this standard metric to the FSWM-HH sample needs to be assessed.

The population of interest in this study of FSWM-HH is hidden and rarely included in global studies that estimate food insecurity. This study’s findings fill a significant gap in the global food security literature and will assist in developing programs to meet and sustain the United Nation’s Sustainable Development Goal 2.2 to diminish hunger among all.

## Methods

### Study design and sample selection

This study data came from a larger mixed-method, population-based, cross-sectional study of the maternal, mental, and nutritional health of FSW who are mothers. Participants were recruited using a purposive sample from urban areas by local sex workers organizations in Kenya and Nigeria and by non-governmental organizations (NGOs) that provide HIV services to FSWs in the DRC. The selection of cities in each country was guided by a study conducted in 2019 by the same research team that established trust among local partners providing services to FSW. Translation of previous study findings into actions implemented by NGOs was the key to gaining trust among FSW. Approximately 100-150 FSWM were recruited in each city. Due to a lack of preliminary data on the main outcome, food insecurity among FSWM-HH, the determination of the sample size was not based on statistical power analysis. Cities included were Nairobi (n=103), Mombasa (n=99), and Kisumu (n=100) in Kenya; Lagos (n=100), Calabar (n=100), and Abuja (n=100) in Nigeria, and Bukavu (n=100) and Kinshasa (n=150) in the DRC. The local NGO partners contacted FSWM in their catchment area and screened those interested in participating for eligibility using the inclusion criteria described below. Case analyses were conducted for those cases deemed incomplete and therefore not included in the analysis.

The study data were collected in January and April 2022. The third wave of COVID-19 ended in 2022 in the region. All lockdown and public health infection control measures were lifted by the time data collection began. Although new COVID variants emerged during the time of data collection, the lockdown measures were not reinstated. The food insecurity measures used collected experiences within the last 12 months starting in January 2022. The COVID-related FIES measurement scale was not used for this data collection.

A representative sample of FSWM-HH was recruited in each city via collaboration with community partners and local sex worker service-providing organizations. A previous study among FSWM by our research team in 2019, used the same framework of study participant recruitment through the same organizations, in the same cities and the sample size we achieved in this study exceeded or was similar (with 72% more recruited in Kenya, 96% in Nigeria and 93% in DRC than in the 2019 study; unpublished data) to the sample size in the previous study. The goal of recruitment was also to have a similar sample size for each city to that of the FAO Gallup World Poll, which identified 100-135 households per cluster [16].

The FSWM participants in the present study met the following inclusion criteria: engaged in full-time sex work for the past three years; age 18 years or older; had at least one biological child age 5 years or younger; and know other FSW. The local partners reviewed the data collection instrument and provided translations during the interviews when necessary. Interviewers were trained and supervised by the lead researcher (BW).

### Ethics approval and informed consent process

The study protocol was approved by the Institutional Review Board of Oregon Health & Sciences University (IRB ID: STUDY0002296) and the Ethics Review Committees in each of the three countries, Kenyata University Centre for Research Ethics and Safety in Kenya, Ecole De Sante Public, University de Kenshasha, DRC and Nigerian Institute of Medical Research, Institutional Review Board, Nigeria. The layperson report summaries were submitted to the local partners before submission to the journal. All eligible participants were informed about the study protocol and their participation was deemed voluntary and they were able to refuse to answer any questions or withdraw at any time during their interview. Interested participants were verbally consented for study participation and indicated consent with an “x” or check mark on the consent form to include FSW who did not read or who could not sign their names. No personal identifiers were collected.

### Data collection and management

Data were collected through individual participant interviews, conducted in Kenya during January and February 2022, in Nigeria during March and April 2022, and in the DRC during February and March 2022. Data were entered into an Excel database and cleaned by one of the data analysts. Further data entry errors such as non numerical entries were corrected by the first author.

#### Survey tool**s**

Items from the World Health Organization (WHO) Demographic Health Survey were used to collect participant demographic information, [19] including age, marital status, and education level. As many FSW live together and often care for children of deceased FSW, information about the total number of adults and children in each household was collected.

The UN Food and Agriculture Organization’s food insecurity experience scale (FIES)was used to collect information about household-level food insecurity experience (Table 1). [14,[16]). The eight-item FIES measures food insecurity indirectly through a series of latent trait questions, rather than directly through a specific question about food insecurity. Three of the eight items are considered subjective measures that ask about being worried about getting enough food to eat, being unable to eat healthy food, and eating less than they thought they should. The other five items are considered objective measures and ask if someone in the household ate fewer foods, skipped meals, ran out of food, suppressed hunger, or did not eat for a full day. The response options to each item are binary (yes/no) with a score of 1 representing an affirmative response and a summative score ranging from 0 to 8. The Rasch analytical model [16] was then used to synthesize individual household-level food insecurity experiences into community or population-level food insecurity experiences along a continuum anchored by mild (starting point) to severe (ending point) food insecurity. The first item of the FIES, “Worried about running out of food because of lack of money,” anchors the household-level continuum at mild food insecurity at the global reference minimum level. The last item of the FIES, “Went without eating one whole day because of lack of money” anchors the household-level continuum at severe food insecurity global reference maximum level. Responses to the other six items, which assess the quality and quantity of food availability, are used to further characterize household-level food insecurity experiences of the community or population between mild and severe at the global reference level [18].

**Table 1:** United Nations Food and Agricultural Organization Eight-Item Food Insecurity Experience Scale.

| Thematic Item | Question | Description[19, 20] |
| --- | --- | --- |
| WORRIED | During the last 12 MONTHS, was there a time when you were worried about you and others in your household would not have enough food to eat because of a lack of money or other resources? | Subjective:<br>Uncertainty and worry |
| HEALTHY | Still thinking about the last 12 MONTHS, was there a time when you and others in your household were unable to eat healthy and nutritious food because of a lack of money or other resources? | Subjective:<br>Inadequate food quality |
| FEWFOOD | Was there a time when you or others in your household ate only a few kinds of foods because of a lack of money or other resources? | Objective:<br>Inadequate food quality |
| SKIPPED | Was there a time when you or others in your household had to skip a meal because there was not enough money or other resources to get food? | Objective:<br>Insufficient food quantity |
| ATELESS | Still thinking about the last 12 MONTHS, was there a time when you or others in your household ate less than you thought you should because of a lack of money or other resources? | Subjective:<br>Insufficient food quantity |
| RANOUT | Was there a time when your household ran out of food because of a lack of money or other resources? | Objective:<br>Insufficient food quantity |
| HUNGRY | Was there a time when you or others in your household were hungry but did not eat because there was not enough money or other resources for food? | Objective:<br>Insufficient food quantity |
| WHOLEDAY | During the last 12 MONTHS, was there a time when you or others in your household went without eating for a whole day because of a lack of money or other resources? | Objective:<br>Insufficient food quantity |

### Statistical analysis

The Rasch model was applied to the food insecurity data analysis [20, 21] such that the latent traits of an individual household, i, item j (j=1 for WORRIED,…j=8 for WHOLEDAY as described in Table 1) is represented as:

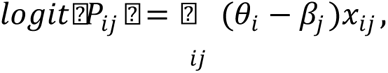

wherein 𝑋_𝑖𝑗_=1 if individual i responded “yes” to item j listed in Table 1, and 𝑋_𝑖𝑗_ = 0, if the response was “no”. The latent trait 𝜃_𝑖_ estimates individual household food insecurity related to the ith person where as 𝛽_𝑗_measures the latent trait corresponding to jth item of the scale.

Data analysis was carried out in two parts. Firstly, given that the FIES has never been applied to the FSWM-HH context, the tool was validated using data gathered from all three countries as a composite. Secondly, detailed analyses of country-level food insecurity experiences were performed across key participant demographic characteristics including, age, marital status, education level, household size (adults), household size (children), HIV status, and pregnancy status. Demographic characteristics evaluated were chosen based on previous research. [6, 22, 23]. The Rasch model R package RM.W was used for FIES analysis and Fisher’s exact test (when counts were less than 5) and chi-square test were used to determine statistically significant within-country demographic differences [20].

FSWM-HH three-country study sample scores were then compared to the FAO global scale. As part of this analysis, the RM.W package skips responses with non-zero values, e.g., a value of 1 representing a response of “yes” to a question by all participants. Therefore, because all participants responded “yes” and received a “1” for their response to FIES Item 1 about being worried they or their household would not have enough food to eat, the scale for WORRIED was obtained through the extended ERM package in the R software,[24] using a randomly assigned negligible amount of zeros approach. This latent trait estimate was only used for visual presentation purposes (Figure 1). As a result, the remaining analyses were carried out using an FAO-certified RM.W package in R software using a seven-item scale excluding Item 1, WORRIED. These item responses are displayed in Figure 1.

**Figure 1:**
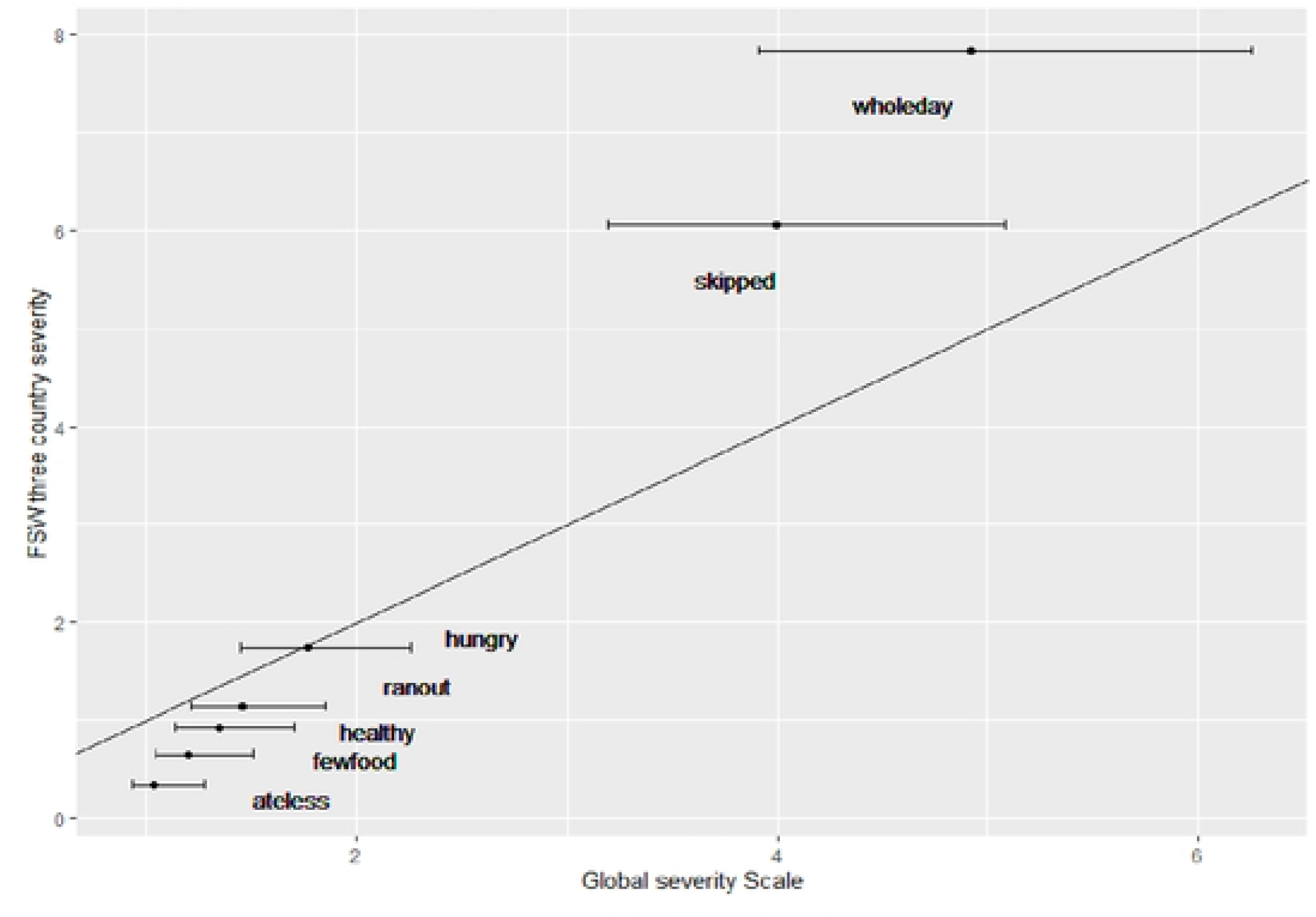
Composite Food Insecurity Experience Scale (FIES) Scores of participants living in the three study countries.

Validation of the FIES for use within the FSWM-HH sample was carried out using 1) reliability estimates of fitting the Rasch model, 2) infit and outfit statistics, and 3) further compared with the global scale by computing comparable estimates after adjusting the three country standards to the global standards, a method described by FAO as calibration. The Rasch model assumption of equal discrimination by each item was tested using infit and outfit statistics to examine outlying values related to each item. Defining a scale of 0.7-1.3 as acceptable for infit statistics, high outfits generally greater than 2 were considered to be unreliable with highly improbable values.[14, 19, 25, 26] The food insecurity measurement fit index measures the Rasch model reliability score and this estimate was used to measure the goodness of fit of the model to the FSWM sample. A fit index Rasch reliability score of 0.7 or above is considered to be a good model fit for the sample of data [15].

To compare country-specific food insecurity scores among FSWM-HH with FAO country-specific estimates, the equating process discussed by the FAO Voices of Hungry project was applied. [4] Specifically, the linear equation below was used to determine the global standard of the three-country metric of item scales (y);

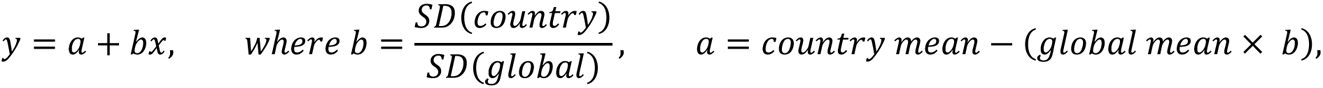

and x is the scaled parameter, item response, for each FIES item. [18]

After rank ordering the severity measure, non-overlapping 95% confidence intervals (CI) were used to decide points of demarcation to classify food insecurity experiences among FSWM-HH as mild, moderate, or severe. Following the direction of the FAO classification, setting the threshold for the classification ranking of mild, moderate, and severe food insecurity was selected as follows. It should be noted that the definition of the latent trait threshold is stated as somewhat arbitrary in their publication [18]. Therefore, we used a statistically determined threshold. The lowest items cluster, designated as “mild”, was selected with the lowest overlapping CI item cluster and there were very few (less than 1%) belonging to this category. The second highest items cluster, designated as “moderate”, was selected with no overlapping confidence intervals to the first cluster but with overlapping confidence intervals with each other. The highest items cluster, designated as “severe”, was selected with the highest-ranking items with overlapping CIs and separated from the other items classified as mild to moderate. Using the Rasch model assigned predictive probabilities and associated CIs, households with the highest probability of severe food insecurity were assigned as experiencing severe food insecurity (HSFI=yes) and the rest was assigned as experiencing mild to moderate food insecurity (HSFI=no). Determinants of HSFI were obtained by fitting a multiple logistic regression model to the dichotomous variable of HSFI. Intersectionality analysis was carried out by fitting logistic regression models, to subgroups identified by the intersections of significant demographic characteristics, in the logistic regression model fit. Our intent was to identify the most vulnerable subgroup for HSFI so that food programs can target them first.

## Results

### Demographic characteristics of FSWM-HH across three study countries

Demographic characteristics of the FSWM-HH sample are shown in Table 2. The majority of participants were in their 20s and were without a partner. The largest percentage of participants in each country had a secondary or higher level of education. The number of adults and number of children in each household varied by country, and the majority of households had one to three children per adult. Noteworthy to mention is that the FSWM participants who indicated having zero children in the household (8.0 % in the DRC, 0.3% in Kenya and 18% in Nigeria) have children living elsewhere and therefore met our inclusion criteria to include only FSW who were mothers.

**Table 2.** Participant Demographic Characteristics.

| **Table 2. Participant Demographic Characteristics** |  |  |  |  |
| --- | --- | --- | --- | --- |
| Characteristic | Democratic Republic<br>of the Congo<br>(n=250)<br>% | Kenya<br>(n=302)<br>% | Nigeria<br>(n=300)<br>% | All three<br>countries<br>(n=852)<br>% |
| <u>Age group (years)**</u> |  |  |  |  |
| 18-20 | 20.4 | 2.0 | 7.7 | 9.4 |
| 21-30 | 75.6 | 54.0 | 62.7 | 63.4 |
| 31-40 | 4.0 | 39.4 | 27.0 | 24.6 |
| 41-52 | 0.0 | 4.6 | 2.7 | 2.6 |
| <u>Marital status**</u> |  |  |  |  |
| Single/never married. | 98.0 | 71.9 | 73.3 | 80.1 |
| Separated/divorced/widowed. | 2.0 | 25.8 | 26.0 | 18.9 |
| Married | 0.0 | 0.7 | 0.0 | 0.2 |
| Not reported | 0.0 | 1.7 | 0.7 | 0.8 |
| <u>Education**</u> |  |  |  |  |
| No schooling | 20.8 | 0.7 | 5.7 | 8.3 |
| Primary | 32.0 | 41.7 | 32.0 | 35.5 |
| Secondary and above | 47.2 | 57.3 | 60.6 | 55.5 |
| Other/Not reported. | 0.0 | 0.3 | 1.7 | 0.7 |
| <u>Household size (adults)**</u> |  |  |  |  |
| 1-3 | 41.6 | 90.1 | 69.7 | 68.7 |
| 4-6 | 37.6 | 8.9 | 23.7 | 22.5 |
| 7 or more | 20.8 | 1.0 | 6.6 | 8.8 |
| <u>Number of children**</u> |  |  |  |  |
| 0 | 8.0 | 0.3 | 18.0 | 8.8 |
| 1-3 | 44.4 | 79.5 | 59.0 | 62.0 |
| 4-6 | 36.8 | 19.5 | 21.3 | 25.2 |
| 7 or more | 10.8 | 0.7 | 1.7 | 4.0 |
| ** p-value for the chi-square test for the difference between country percentage across demographic characteristics < 0.0001 |  |  |  |  |

The demographic profiles of FSWM-HH across the three study countries were different (Table 2) and the participants living in the DRC were demographically different from those living in Nigeria and Kenya. FSWM-HH in the DRC were statistically significantly younger, were single or never married, had achieved a lower education level, had larger sized households, and had a higher percentage of households with more than 3 children compared to FSWM-HH in Kenya or Nigeria (p<0.0001).

Participants’ clinical and health behavior characteristics frequency distribution differed by country (Table 3). The majority of participants did not have or had not tested for HIV, and the majority were not pregnant at the time of survey administration. Across all three countries, 96% of participants who reported being HIV-positive were on ART. Participants living in Nigeria had the highest rate of HIV, followed by those living in the DRC and Kenya. Participants living in Kenya had the lowest percentage of FSWM who were currently pregnant. The majority of participants reported consuming alcohol during a pregnancy with a large percentage reporting substance use during a pregnancy. Differences in clinical characteristics of participants across countries were statistically significant (p<0.0001).

**Table 3:** Participant Clinical and Health Behaviour Characteristics.

| **Table 3: Participant Clinical and Health Behaviour Characteristics** |  |  |  |  |
| --- | --- | --- | --- | --- |
| Characteristic | DRC<br>(n=250)<br>% | Kenya<br>(n=302)<br>% | Nigeria<br>(n=300)<br>% | All three<br>countries<br>(n=852)<br>% |
| <u>HIV status**</u> |  |  |  |  |
| Negative | 48.0 | 75.8 | 79.0 | 68.7 |
| Positive | 5.6 | 16.9 | 8.3 | 10.6 |
| Not reported/not tested | 46.4 | 7.3 | 12.7 | 1.5 |
| <u>Pregnant **</u> |  |  |  |  |
| Yes | 17.6 | 3.0 | 16.3 | 12.0 |
| No | 82.4 | 95.7 | 82.7 | 87.2 |
| Not reported | 0.0 | 1.3 | 1.0 | 0.8 |
| <u>Alcohol use during any pregnancy**</u> |  |  |  |  |
| Yes | 77.2 | 52.3 | 72.0 | 66.5 |
| No | 19.6 | 44.7 | 26.6 | 31.0 |
| Not reported | 3.2 | 3.0 | 1.4 | 2.5 |
| <u>Substance use during any pregnancy**</u> |  |  |  |  |
| Yes | 62.0 | 29.1 | 18.3 | 35.0 |
| No | 34.0 | 67.5 | 79.0 | 61.7 |
| Not reported | 4.0 | 3.4 | 2.7 | 3.3 |
| ** statistically significant for the chi-square test for the difference between country percentages with $p\text{-value} < 0.0001$ | | | | |

### Validity of the FIES tool for use among samples of FSWM in the three study countries

Results of the responses to the FSWM-HH FIES analysis in the DRC, Kenya and Nigeria are shown in Figure 1, in the composite food insecurity reference continuum. All study participants were worried about not having enough food to eat which was the anchoring measurement for the “mild” food insecurity side of the continuum with a score of 0.05 (95% CI:0.01-0.4), which was obtained from ERM package in R software [24]. Due to the lack of inter-individual variability, this item was excluded from the item severity and individual severity estimations using the FAO-recommended RM.W package. “WHOLEDAY” (not eating for the whole day) had the highest score of 7.8 (95% CI: (5.9-10.4) and was defined as the anchoring measurement for the “severe” side of the food insecurity continuum for both groups in the comparison. Participants who were referenced as skipping meals with a score of 6.1 (95% CI: 4.5-8.2) (“SKIPPED” in Figure 1) were grouped into the severe food insecurity group, due to overlapping 95% CI with “WHOLEDAY”, as opposed to classification in the global reference scale as moderate food insecurity [17]. The negative small sample correlation (r=-0.03, p=0.36) between the responses to these two items, “SKIPPED” and “WHOLEDAY”, were statistically independent and indicates that most FSWM-HH in the severe food insecurity group use either skipping a meal or not to eating for the whole day to address food insecurity. Similar to the global scale, our moderate food insecurity categorization was determined by the two items of hungry and running out of food (“HUNGRY”, “RANOUT” in Figure 1). However, the FSWM-HH scale classification of inability to eat healthy foods (“HEALTHY” in Figure 1) also fell into the moderate food insecurity classification compared to being in the mild classification on the global scale. Overall, FIES item classification of FSWM-HH was similar to the global scale with similar item classification of all three categories (mild, moderate, and severe) for 5 of the 8 survey items.

The fit of the Rasch model was assessed by using the fit index score, infit and outfit statistics. All infit statistics were within the range of 0.82 (for “RANOUT”) to 1.1 (“HEALTHY”). In the literature, infit values within the range of 0.7 to 1.3 are considered to justify the Rasch model assumption of equal discrimination [27]. In addition, the model fit index reliability score of 0.75 was deemed a good model fit given this value exceeded 0.7, the lower bound of the acceptable range suggesting the estimated item responses are reliable estimates of food insecurity for the sample of FSWM-HH.

### Calibration of FSWM-HH Scale using Global Reference Standards

To compare prevalence across subgroups, it is recommended that FIES item severity scores be calibrated using Global 147 Country Item Scales as the reference [18, 20]. The results of calibration against the global metric are shown in Table 4. Based on FAO instructions on FIES tool use, when calibrating and equating, outliers should be excluded.[28] Therefore, the two items “WHOLEDAY” and “SKIPPED”, the two major outliers of the item score distribution, were omitted in the calibration equating linear interpolation parameters (intercept and slope) calculation [28]. Nevertheless, the adjusted scores shown in Table 4, show the linear interpolation for these two items after calculating the intercept and the slope based on the included items.

**Table 4:** Three Country Female Sex Worker-Mother Head of Household and Global 147 Country Thematic Item Severity Scale Values.

|  |  |  |  |
| --- | --- | --- | --- |
| Thematic Item | Three-Country 7-Item Severity Score<sup>a,b</sup> | Global 147 country reference scale-7-Item Severity-Score<sup>**</sup> | Global Standard calibrated on Three-Country Metric |
| HEALTHY | 0.93 (0.53,1.62) | -0.85 | 1.35 |
| FEWFOOD | 0.65 (0.34,1.25) | -1.11 | 1.21 |
| SKIPPED | 6.05 (4.50,8.15) | 0.35 | 4.0<br>(adjusted threshold)<sup>c</sup> |
| ATELESS | 0.34 (0.14,0.79) | -0.31 | 1.0 |
| RANOUT | 1.14 (0.68,1.91) | 0.51 | 1.5 |
| HUNGRY | 1.74 (1.12,2.69) | 0.75* | 1.8 |
| WHOLEDAY | 7.83 (5.89,10.42) | 1.88 | 4.9<br>(adjusted threshold)<sup>c</sup> |
| Mean | 2.67 | 0.1740643* | 2.247913 |
| SD | 2.99 | 1.023484 | 1.548293 |

<sup>a</sup> 7-Item Severity Score was calculated after excluding responses for “WORRIED”
<sup>b</sup> Mean (95% Confidence Interval)
<sup>c</sup> values were calculated using the mean and SD (standard deviation) calculated without the items (extreme values, see below).
\*Source:
(https://www.fao.org/fileadmin/user_upload/voices_of_the_hungry/docs/Cafiero_Nord_Viviani_IMEKO_2016_rev_2.pdf)
\*\*FAO statistics downloaded from https://www.fao.org/faostat/en/#data/FS

When reference scales are made on a common scale, the two objective measures of skipping meals (“SKIPPED”) and not eating for a whole day (“WHOLEDAY”) due to lack of money and resources for the FSWM-HH sample were discriminant and exceeded the Global Reference Scale metric (Figure 2). The other objective measure of “HUNGRY”, interpreted as “did not eat when hungry” was similar to the Global Reference Scale. The subjective measure of “ATELESS”, “eating less and thinking not enough” item received the lowest severity score on the scale. Very few FSWM-HH participants (n=8, 0.9%) were classified as mildly food insecure, 22.1% were classified as moderately food insecure and 77% were classified as severely food insecure. Further analysis was carried out using two classifications of mild to moderate combined (noted as moderate onwards) and severe.

**Figure 2:**
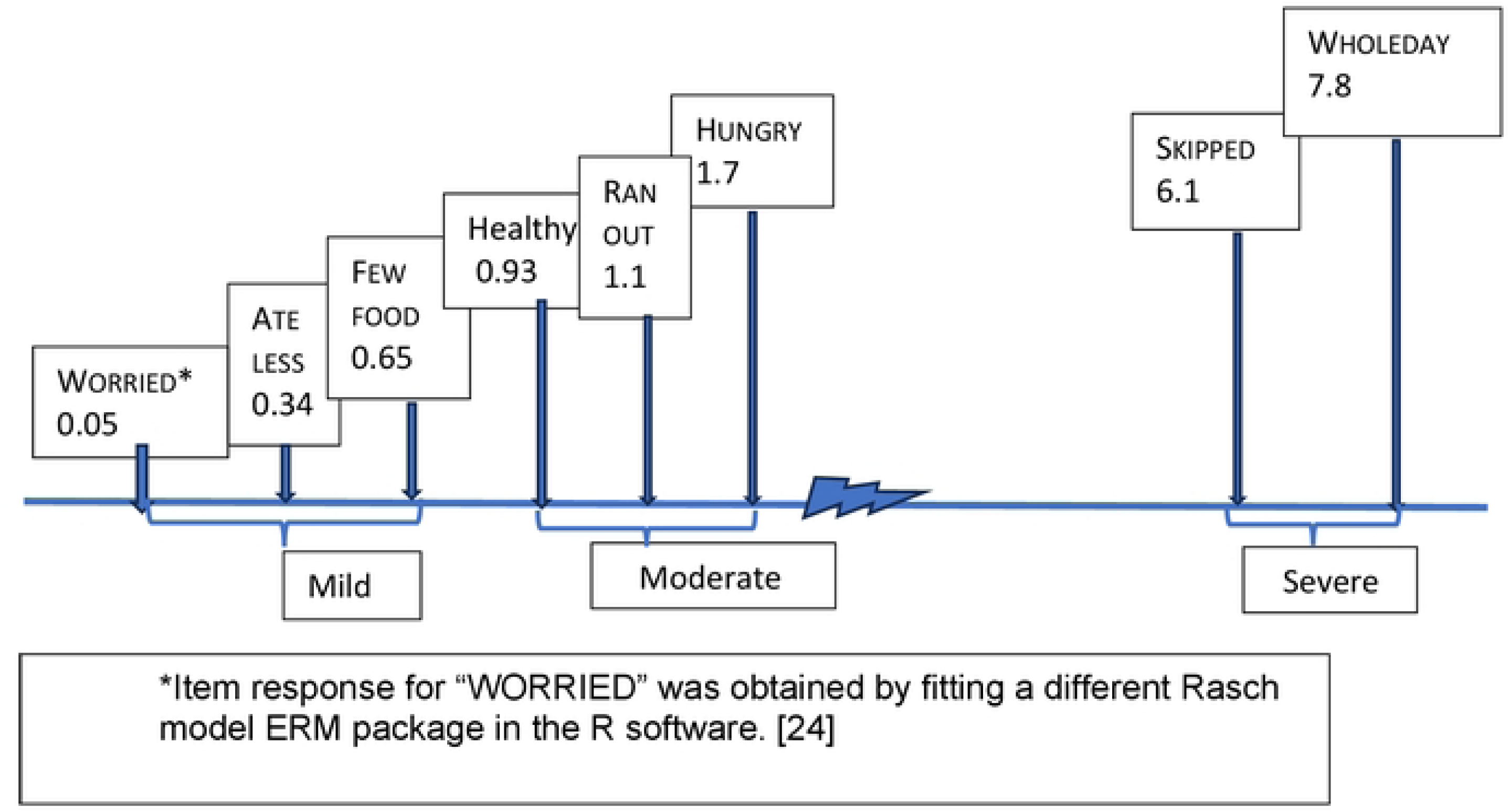
**Relationship between 3-country FSWM-HH and Global 147-country Food Insecurity Severity Scales, after calibration into the same scale.**

### Comparison of FSW-MHH food insecurity prevalence with country-level estimates

Country-level food insecurity prevalence among FSWM-HH was calculated by the level of household food insecurity severity, classified as moderate and severe, using individual parameter estimates. The percentage was calculated using the total country FSWM-HH sample size as the denominator (Table 5). The FAO country-level estimates (Table 5) came from the FAO statistics estimates derived from a “*nationally representative samples of the adult population (aged 15 and older) in each country*”[18]. The demographic characteristics of the FAO sample in the Sub-Saharan African region differ from that of the study sample of this paper [26]. In the DRC, Kenya, and Nigeria, the prevalence of severe food insecurity is significantly higher among FSWM-HH compared to FAO country-level estimates for the same country. Conversely, the prevalence of moderate food insecurity is significantly lower among FSWM-HH compared to country-level estimates for the same. If the country-level FAO food insecurity survey sample did not include FSWM-HH, then we can assume that the prevalence of severe food insecurity among FSWM-HH is 1.5 times higher in DRC, nearly 3 times higher in Kenya, and 4.5 times higher in Nigeria as the prevalence of severe food insecurity within the rest of the country population.

**Table 5:** Prevalence of Moderate and Severe Food Insecurity among the Three-country Female Sex Worker Sample and the FAO 147-Country Estimates.

|  |  |  |  |  |
| --- | --- | --- | --- | --- |
| Country | Prevalence of Food Insecurity by Severity among the Three Country FSWM-HH Sample (95% CI)** |  | Prevalence of Food Insecurity by Severity among the FAO Country Specific Community Sample* |  |
|  | Moderate | Severe | Moderate | Severe |
| DRC | 40.4<br>(34.3-46.8) | 59.6<br>(53.2-65.7) | 69.2<br>(66.9-71.5) | 38.5<br>(36.1-40.9) |
| Kenya | 22.2<br>(17.6-27.3) | 77.8<br>(72.7-82.4) | 69.5<br>(67.9-71.1) | 26.1<br>(25.0-27.1) |
| Nigeria | 11.0<br>(7.70-15.1) | 89.0<br>(84.9-92.3) | 62.9<br>(60.1-65.8) | 19.8<br>(17.6-21.9) |
| All three countries | 23.6<br>(20.7-26.6) | 76.4<br>(73.4-79.2) | -- | -- |
| ** Exact (95% Confidence Interval) |  |  |  |  |
| *Source: Three-year average 2018-2020 estimates from FAO stat data<br>https://www.fao.org/faostat/en/#data/FS |  |  |  |  |

These within and cross-country differences in the prevalence of severe food insecurity among FSWM-HH are statistically significant (p<0.0001).

### Household severe food insecurity (HSFI) prevalence across individual and household demographic characteristics

The prevalence of severe food insecurity across the head of the household demographic characteristics, related to a single determinant impacting HSFI when unadjusted for other determinants, are listed in Table 6. Of the demographic characteristics of the head of the FSWM-HH considered, older age, living without a partner (unmarried, widowed, or separated), having no schooling or only primary schooling, and having 1-3 children living in the household significantly determined having higher HSFI in the three-country FSWM-HH sample.

**Table 6:** Single determinant of severe food insecurity in the three study countries.

|  |  |  |  |  |
| --- | --- | --- | --- | --- |
| Characteristic | DRC<br>Severe FI<br>% | Kenya<br>Severe FI<br>% | Nigeria<br>Severe FI<br>% | All three<br>countries<br>Severe FI % |
| Age group (years) |  |  |  |  |
| 18-20 | 66.7 | 83.3 | 95.7 | **76.3** |
| 21-30 | 56.6 | 75.5 | 87.8 | **73.2** |
| 31-40 | 80.0 | 79.8 | 88.9 | **83.3** |
| 41-52 | NA | 85.7 | 100.0 | **90.9** |
| p-value* | 0.20 | 0.80 | 0.79 | 0.01 |
| Marital status |  |  |  |  |
| Single/never married. | 60.4 | 76.0 | 88.0 | **74.2** |
| Separated/divorced/widowed. | 20.0 | 83.3 | 92.3 | **85.7** |
| Married | NA | 50.0 | NA | **50.0** |
| Not reported | NA | 80.0 | 100 | **85.7** |
| p-value* | 0.16 | 0.30 | 0.4 | 0.005 |
| Education |  |  |  |  |
| No schooling | 69.2 | 50.0 | **100.0** | **76.1** |
| Primary | 62.5 | 84.1 | **96.9** | **82.5** |
| Secondary and above | 53.4 | 73.4 | **83.5** | **72.3** |
| Other/Not reported | NA | 100.0 | **100.0** | **100.0** |
| p-value* | 0.12 | 0.05 | 0.002 | 0.006 |
| Household size (adults) |  |  |  |  |
| 1-3 | **51.0** | 78.3 | 91.4 | 78.1 |
| 4-6 | **61.7** | 74.1 | 83.1 | 71.4 |
| 7 or more | **73.1** | 66.7 | 85.0 | 76.0 |
| p-value* | 0.02 | 0.60 | 0.10 | 0.16 |
| Number of children |  |  |  |  |
| 0 | **70.0** | 0.0 | 88.9 | **82.7** |
| 1-3 | **70.3** | 76.3 | 88.1 | **79.0** |
| 4-6 | **46.7** | 84.7 | 90.6 | **70.2** |
| 7 or more | **51.8** | 100.0 | 100.0 | **61.8** |
| p-value* | 0.004 | 0.13 | 0.91 | 0.006 |
| HIV status |  |  |  |  |
| Not Tested | **73.0** | 82.4 | 80.6 | 75.5 |
| Negative | **50.0** | 75.1 | 91.1 | 76.5 |
| Positive | **28.6** | 86.3 | 80.0 | 75.6 |
| Not reported | **100.0** | 100.0 | 85.7 | 92.3 |
| p-value* | 0.0001 | 0.25 | 0.08 | 0.86 |
| Pregnant |  |  |  |  |
| Yes | 52.3 | 88.9 | 89.8 | 73.5 |
| No | 61.2 | 77.8 | 88.7 | 76.9 |
| Not reported/Did not know | 0.0 | 50.0 | 100.0 | 71.4 |
| p-value* | 0.36 | 0.32 | 1.0 | 0.63 |
| Alcohol use** |  |  |  |  |
| Yes | 58.6 | 81.6 | **86.1** | 75.4 |
| No | 69.4 | 72.6 | **96.3** | 79.2 |
| Not reported | 25.0 | 88.9 | **100.0** | 66.7 |
| p-value | 0.05 | 0.13 | 0.02 | 0.29 |
| Substance use** |  |  |  |  |
| Yes | 62.0 | 80.7 | 85.5 | **71.8** |
| No | 58.8 | 76.5 | 89.5 | **79.5** |
| Not reported | 30.0 | 80.0 | 100.0 | **67.9** |
| p-value | 0.15 | 0.81 | 0.53 | 0.02 |
| *p-value reflects statistically significant differences across age groups for the Fishers exact test to account for small cell size, otherwise, the two-sample proportion test p-values are reported.<br>** during pregnancy. |  |  |  |  |

There was a significant trend with older FSWM-headed households showing a higher prevalence of HSFI although this socio-ecological relationship did not reach statistical significance when stratified by country (Table 6). In the DRC (p=0.12) and Nigeria (p=0.002), the heads of households with no schooling and in Kenya those with a primary education level (p=0.05) had a higher prevalence of HSFI, than those with secondary or above levels of education. Household size showed a significant difference to food insecurity only in the DRC where households with seven or more members had the highest prevalence of HSFI. In the DRC, having no or 1-3 children in a household significantly determined a higher prevalence of HSFI, and those FSWM-HH with heads who had never tested for HIV had the highest prevalence of HSFI. Country-level HSFI prevalence did not change when classified by FSWM-HH heads’ pregnancy status, or substance and alcohol use during pregnancy except in Nigeria. In Nigeria, those heads who stated they did not use alcohol during pregnancy had a significantly higher prevalence of HSFI than those households whose head did use alcohol during pregnancy (p=0.02). When all three country responses related to substance use were combined those heads who stated they did not use substances during pregnancy had a higher prevalence of HSFI (p=0.02).

### Determinants of severe food insecurity: Multivariable analysis results

Multiple logistic regression results revealed the adjusted odds ratio (AOR) of household severe food insecurity when multiple factors simultaneously existed in households. In the three-country combined sample, those FSWM-HH having no children living with them (adjusted odds ratio=AOR=2.3, 95% CI: 1.14-4.45) or 1-3 children living with them (AOR=1.63, 95% CI: 1.13-2.37), compared to those having 4 or more children living with them, were more likely to experience HSFI although this trend was only statistically significant for FSWM-HH in the DRC. In the three-country combined sample, those FSWM-HH with heads having a lower education level (primary education) were twice as likely to be severely food insecure compared to those households with heads having a secondary education level (AOR=1.92, 95% CI:1.33-2.78). This trend was statistically significant in Nigeria where FSWM-HH with primary education levels were more likely to be categorized as having HSFI (AOR=6.4, 95% CI: 1.90-21.8). Overall, those households with FSWM heads who did not use substances during pregnancy were more likely to be classified as having HSFI (AOR=1.45, 95% CI: 1.03-2.04) compared to households with substance-using FSWM heads. This trend was marginally significant.

### The HSFI at the Intersection of Socio-demographic Vulnerability Indicators

In this section, we present the prevalence of severe food insecurity at the intersection of demographic factors that were significant in the single-factor analysis shown in Table 6. This analysis is limited to the combined sample, in most cases, since there were not enough data to carry out individual country-level analyses in some countries. Four intersections, with salient demographic characteristics informed by the aforementioned analysis results, were determined to include the number of children in the household, living with or without a partner, and the level of education of the FSWM-HH.

Firstly, we examined the impact of age of the FSW head of the household with no partner, since the majority had no partner. Among the no-partner-headed households, younger FSWM <u>(<</u> 30 years of age) prevalence of HSFI was 83.2% (10.4% lower with 95% CI: 4.2% to 16.7%) compared to households headed by older FSWM (above 30 years of age) with no partners (p=0.002). Secondly, we compared the socio-ecological differences of having four or more versus fewer than four children in a household within the group of households with the lowest level of education, identified as no schooling or primary education. In this group of households headed by FSWM with primary or no schooling, those with 0-3 children in the household had a prevalence of HSFI of 85.0% (p=0.03), which was 10% higher than (95% CI: 0.3% to 19%) the households headed by FSWM with four or more children. Thirdly, we compared the impact of having fewer or more children in the household (0-3 children compared to 4 or more children) among FSWM heads group with secondary or higher education. In this group of households, those with 0-3 children had an HSFI prevalence of 76.0%, which was 13% higher (95% CI: 3% to 23%, p=0.008) than those with four or more children. Fourthly, we compared the prevalence of HSFI between two of the most vulnerable groups of households with two combined demographic identities of FSWM heads, having 0-3 children in the household and with primary or no schooling to households headed by FSWM and compared with the FSWM-HH having four or more children in the household and secondary or higher education. This intersectionality analysis revealed the most vulnerable group for severe food insecurity was FSWM-HH with 0-3 children in the household with no schooling or primary education which had a prevalence of a HSFI of 85%. The prevalence of HSFI in this subgroup was 22% higher (95% CI: 12%-32%, p=0.00001) than the prevalence of HSFI in the comparison subgroup of FSWM-HH with four or more 4 children in the household and a head with secondary or higher education level.

## Discussion

Rasch modeling was used to estimate the prevalence of severe food insecurity among FSWM-HH in three LMIC in Sub-Saharan Africa. The FIES tool has not been specifically or exclusively applied to the FSWM-HH population before and the analyses presented here validate the tool for the use in this vulnerable population, through the exclusion of one item related to being worried about not having food and also calibrated against the global scale. Moreover, the model reliability was at the accepted level for a good fit and all assumptions for the model fit were met. Some caveats noted were the degree to which responses were homogeneous reducing the inter-subject variation. For example, the majority of FSWM, 649 out of 852, answered yes to all items in the FIES indicating severe insecurity, and all 852 answered yes to being worried about food. Given that every FSWM was worried about not having enough food to eat, we omitted this item from the model and used a seven-item scale to measure food insecurity among FSWM-HH. Nevertheless, our Rasch model fit reached the required level of model fit statistic overall (Fit Index=0.75), and item infit, which detected outliers, extreme values, and outfit statistics which detected item response discrimination were all within the required limits. The infit statistics (assessment of model fit) were below the values noted in the literature for Nigeria except for “SKIPPED” in which our infit value was 1.1 and the Nigeria community value noted from the WHO assessment was 1.0 [26]. Similarly, our outfit statistics for all items were below the WHO assessment for Nigeria noted as 0.1[26]. This comparison further validates our Rasch model fit to the FSWM-headed household food insecurity assessment.

The FSWM-HH sample estimated household food insecurity item scores were calibrated using the global scale and then compared with the FAO country-level estimates. The anchoring points of severe food insecurity in the study sample were marked by the two items skipping meals (SKIPPED) and not eating for the whole day (WHOLEDAY). The prevalence of severe food insecurity among FSWM-HH as estimated in the DRC, Kenya, and Nigeria is 1.5, 3, and 4.5 times as high, respectively, as the country-level FAO estimates for the general population. In particular, FSWM-HH in Nigeria and Kenya were skipping meals, not eating for the whole day, and suppressing hunger, because they did not have enough money to buy food or lacked resources. Although the situation for FSWM-HH in the DRC was less severe, HSFI prevalence was nonetheless 1.5 times the FAO estimated country-level prevalence. The country-level prevalence was estimated using a lower cut-off of having answered yes to 7 or more items [26] and we used a higher cut-off of 8. The severe food insecurity situation among FSWM-HH would be even worse if the same cut-off of 7 affirmative responses had been used.

Determinants of HSFI among FSWM-HH were the same as in the literature with one exception of the composition of children. Previous research findings suggest food insecurity is positively associated with sexual risk behavior and poor pregnancy outcomes in women, [8] in addition to living in low-income households [29]. A study conducted in Nigeria with a sample of 91.2% female households indicated a 66% prevalence of food insecurity [30]. Even further, a study conducted in Sub-Saharan Africa confirmed a higher prevalence of food insecurity among female-headed households [31]. All of these common country and gender-level associated risks may contribute to the high prevalence of severe food insecurity estimated in our FSWM-HH study sample.

Our sample’s HSFI experiences differ in magnitude by household characteristics noted in the literature for the general population in LMICs. The estimated HSFI prevalence in our study sample was higher among headed by older FSWM (aged between 31-52 years) than younger FSWM headed (30 years or less) households. This trend is similar in the general population global statistics among developing countries [29]. However, age is noted as an insignificant predictor of food insecurity in developed nations [32]. Consistently across all three study countries, our sample estimated prevalence of FSWM-HH HSFI was higher for those heads living without a partner which is consistent with global research studies among developing countries [32].The direction of HSFI trend across FSWM-HH heads’ level of education was also similar to that reported in South Africa and East Africa, but the magnitude of severity in prevalence among our sample in Nigeria was three times higher odds of household severe food insecurity compared to the odds reported for East Africa [33], though similar to the odds reported for South Africa [31]. This evidence from studies in the same region affirms our findings that those households with lower education level heads, experience higher odds of HSFI. Concerningly, we also found that households with fewer children living in the household, 0-3 children, as having higher odds of HSFI than households having four or more children. This trend is counterintuitive and different than reported in other Sub-Saharan countries, in which households with four or more children found to have higher odds of HSFI [26]. Our data suggest that the age range of households with four or more children varies between 18 months to 16 years. These older children may be contributing to securing food or getting food elsewhere and also help to take care of younger siblings, allowing adult members to engage in income generating activities.

Our findings suggest FSWM-HH share some of the structural-demographic determinants of severe household food insecurity as the general population within the country or region, but the magnitude of the prevalence is much higher among our study population. Moreover, our intersectionality analysis identified the highest severe food insecurity group was headed by FSWM-HH with 0-3 children in the household, and those who had no schooling or primary education, a finding which is consistent across all three study country subgroups of FSWM-HH.

### Limitations and future directions

Many studies have found income as the strongest determinant of severe household food insecurity [8, 26, 29, 34, 35]. However, we did not collect data on household income, which may be highly variant within and between FSWM-HH. Sample characteristics and prevalence of these characteristics were different across study countries and may be attributed to sample recruitment strategies used in the DCR where participants were recruited and data were collected by HIV service-providing NGOs, not local sex workers organizations as in Kenya and Nigeria.

Future studies should assess the types of food consumed by these households, which family members skip meals and go without food for an entire day, demographic characteristics of household members, both children and adults, and household income (per diem per adult), and available resources. In addition, future studies of food insecurity among FSWM-HH should recruit and evaluate larger samples to generate greater inter-person/inter-household variability.

## Conclusion

The FAO FIES scale can be used to measure household food insecurity among the extremely vulnerable population of FSWM-HH living in LMIC in Africa. We found that in each country studied, the prevalence of severe food insecurity among FSWM households was 1.5, 3, and 4.5 times higher than the country-level prevalence of severe food insecurity noted by the FAO using the 2018-2021 country average estimates. In the population of FSWM-HH that we studied, skipping meals and not eating for an entire day reflected the level of severe food insecurity experienced. Demographic characteristics of the study sample household heads were different across the three countries evaluated; however collectively, older age, lower level of education, heading a household without a partner, and having fewer (0-3) children living in the household were strong determinants of household severe food insecurity. Our analyses identified the most vulnerable FSWM-HH subgroup of severe food insecurity have 0-3 children living in the household, headed by FSWM without a partner, have no schooling or a primary education, and are older than 30 years. These findings could be immediately used to design food programs and welfare policies that reduce severe food insecurity among these households. Specifically, although FSWM-HH with 0-3 children are at higher risk for severe food insecurity, all children regardless of household size need protection against malnourishment. Sufficient funding is urgently needed to mitigate HSFI among the most vulnerable mothers and children in the world. Global food programs should recognize the unique situation of and needs of FSWM and their children and target food relief to them. Also monitoring food insecurity among this very vulnerable and marginalized population of women and their children should be prioritized.

## Data Availability

The senior author, BW, affirms that the manuscript is an honest, accurate, and transparent account of the data collected, stored, and analyzed. Due to ethical considerations (i.e., consent was not obtained or given for open data access or additional data usage), controlled and secure data access and usage is necessary for subject protections. De-identified aggregate data used for this analysis can be requested from Brian Willis at. Access permission will be considered based on the following usage criteria: (a) for the purpose of partnering on research on female sex workers (b) for inclusion in curriculum for educational purposes or (c) for the provision of services to female sex workers and their children by governmental and non-governmental organizations.

## Acknowledgments

We would like to thank all women who participated in this study and our local partners including: Coast Sex Workers Alliance (Mombasa, Kenya); Kisumu Sex Workers Alliance (Kisumu, Kenya); SWOP Ambassadors (Nairobi, Kenya); cPHDP Champions (Nairobi, Kenya); Kiambu Sex Workers Alliance (Thaki, Kenya); Bar Hostess Empowerment and Support Programme (Nairobi, Kenya); Royal Women’s Health and Rights Initiative (Lagos, Nigeria); Initiative for Young Women’s Heath and Development (Calabar, Nigeria); Nigerian Sex Workers Alliance, also know as Precious Diamonds (Abuja, Nigeria); Parlons SIDA aux Communautaires (Kinshasa, DRC); Cadre De Recuperation Et D’encadrement Pour L’epanouissement Integral Des Jeunes (Kinshasa, DRC); Association Pour Le Soutien, L’ Education, La Promotion De La Vie Et des Initiatives Communautaire (Kinshasa, DRC) and Action Humanitaire pour la Santé et le Développement Communautaire (Bukavu, DRC).

## Author contributions

Swarna D.S. Weerasinghe contributed to the conceptualization, data management, analysis, interpretation of results and writing the manuscript. Brian Wills, secured funding, designed the study, trained and supervised local staff in the data collection, coordinated data entry and cleaning, and contributed to writing and reviewing the manuscript. Jennifer A. Jackson, Diane D. Stadler and Wendy L. Macias-Konstantopoulos, contributed to reviewing and editing the manuscript.

